# Trust and transfer during the covid-19 pandemic: did digital cash transfer save lives?

**DOI:** 10.1101/2024.04.04.24305360

**Authors:** Gindo Tampubolon

## Abstract

**BACKGROUND:** In the first semester of 2020 one in six people in the world (1.36 billion) received cash transfers to tide them over the spreading pandemic that originated in Wuhan. By December 2021 it had claimed up to 18.2 million excess deaths. Compared to no (digital) transfer, did digital cash transfer reduce excess deaths? Serendipitously, two years earlier the world reported levels of trust in science. Did such trust inoculate societies from the pandemic?

**MATERIALS & METHODS:** The growing excess deaths literature distinguishes causal factors (e.g. digital transfer) from risk factors (e.g. trust). During the pandemic period, no randomised trials of digital transfer with excess deaths as primary outcome were registered. This study used reports from 170 countries and applied endogenous treatment models to overcome the endogeneity of digital transfer.

**RESULTS & DISCUSSION:** I found that serendipity matters: countries with high trust in science suffered fewer excess deaths. But creativity matters more. Digital transfers –some creatively scrambled from scratch– reduced excess deaths by many more. Equally marked, North-South inequity in excess deaths persists, consistent with the concentration of vaccine distributions in the North early on. All three are statistically significant.

A series of robustness analyses points to the results being reliable to change in outcome estimates, change in trust sources, and change in treatment of omitted countries. Mechanistic analyses show evidence that digital transfer created leg room for governments to expand stringent restrictions to control the spread of SARS-CoV-2, while in the South it weakened the correlation between informal economy and excess deaths. This study of the causal effect of digital cash transfer on a hard outcome (excess deaths) revealed ample global digital dividends across the largest number of countries. This new evidence also suggests that improving and monitoring trust in science can offer considerable benefits for humanity.

## INTRODUCTION

On 5 May 2023 the World Health Organization (WHO) lifted the global emergency of coronavirus 2019 disease (covid-19), three years after announcing the emergency. The covid-19 pandemic, originating in Wuhan China, was confirmed to have claimed nearly seven million lives. Shortly afterwards the WHO released updated numbers of excess deaths in 2020-2021 (14.9 millions), critical to any effort to understand the causes and consequences of this century-timescale tragedy which wrought immense suffering in people’s lives and livelihoods.^1,2^ The early months of the pandemic witnessed an unprepared world under threat.

Broadcast and broadsheet media and especially social media shared images of the world caught unaware, of health systems overwhelmed, of three-tiered graves outside big cities with noisy earth diggers stacking coffins *sans* rites, and of caretakers in white disposable personal protective equipment. That the world was caught unaware does not mean no information at all was ready for learning from the episode. Government responses to the pandemic have been tallied by international organisations such as the World Bank on cash transfer or social protection, the Johns Hopkins University on cases and deaths across the world, and Oxford University on government restrictions.^3–7^ All this simply means creative use of patchy information is imperative to garner lessons from the covid-19 pandemic.

Within six months in 2020 governments and donors had sprung into action deploying drastic responses such as social restrictions and financial support, transferring cash to one in six people around the world.^6^ Some governments devised cash transfer or payment programmes to support people in lockdown, in furlough or in unemployment, some of which were delivered as digital cash while others as non-digital cash. Until now it is not clear whether digital cash transfer (compared to non-digital or no transfer altogether) led to fewer excess deaths. Estimating the causal effect of the digital cash transfer is important because such programmes are vast. And it can inform debates on global digital dividends. Without scrutiny of such programmes there is little way of knowing whether the tens of millions of excess deaths were followed by billions of misplaced funds.

The scale of excess deaths around the world due to the covid-19 pandemic was immense, between 14.9 to 18.2 million deaths. Such magnitudes quickly invited attempts at explanation.^8,9^ The resulting literature explaining both excess deaths and direct deaths from covid-19 has identified various kinds of trust as important risk factors. Drawing from the Wellcome Trust Global Monitor (second) survey in 2020 and other global sources, Sun and colleagues found that six important factors could explain excess deaths in 80 countries, including trust in government pandemic advice, vaccine coverage by November 2021, confidence in hospitals, average income per person, adult obesity rate and older population rate.^10^ Focusing on a different kind of trust, Ji and colleagues identified trust in government or politics as an important factor, negatively associated with log of covid-19 deaths.^11^ The trust variable is a latent factor derived using Bayesian methods from responses to trust in government questions asked in 10 world surveys fielded between 1990 and 2019. The estimates for 134 countries in 2019 are used. Both studies found that more trust is associated with fewer deaths. Crucially, the literature begins to distinguish factor from cause or association from effect. This new work adopts this distinction and estimates both the association with trust in science in 2018 and the effect of digital cash transfer.

In following this literature two modifications are made. First instead of the trust in government advice during the 2020 pandemic or trust in government in 2019, I use trust in science in 2018 from the Wellcome Global Monitor.^12^ The timing is chosen to avoid reverse causality. Second, the object of trust viz. science instead of government is chosen because of the global nature and scientific nature of the threat. For global threats such as pandemic and planetary climate change, science is deemed important and trust in science (as distinct from the science) is equally crucial. This work also includes the global north-south distinction because it carries consequences not least in vaccines development and coverage. Vaccines were of course the essential requirement for the world to emerge from the pandemic to return to normal ways of life, and these were unequally distributed across the global divide.^13–15^

This work contributes to the fundamental role of trust in science when tackling global shocks and global secular (long term) threats. It also speaks to a practical response when faced with global shocks. Trust and digital transfer are evidently critical. Trust in science is indispensable because the virus was novel and has not been known to humanity and has been mutating rapidly even as the reservoir of infected people increased and fluctuated, proving to be a fertile breeding ground for new variants of interest to emerge through sheer random mutations. The novelty of the threat was its defining feature that required science to step up. That made trust in science critical. The link novelty - science - trust in science is immediate and crucial.

As a case in contra point, the American scientist Oreskes,^16^ author of *Why Trust Science*, opened with the American response to the pandemic (:ix),

> Covid-19. Rarely does the world offer proof of an academic argument, and even more rarely in a single word. But there it is. Covid-19 has shown us in the starkest terms - life and death - what happens when we don’t trust science.

Contrast this quote with a riposte from a senior UK government minister, Michael Gove reported in *The Financial Times* with regards to Britain’s exit from the European Union (FT 13 June 2016. ‘Britain has had enough of experts, says Gove’).^17^

Like Oreskes’ words, this work speaks to another global secular threat: climate change. Trust in science is a matter of life and death for a person or the planet. This work points towards steps that can be taken to mount an effective response to the threat of climate change, namely improving trust in science and monitoring such trust. It is shown here that the instrument used is simple (following the literature).^18,19^ Even so it has external validity – an essential quality of a scientific concept – showing that the instrument is significantly related to covid-19 excess deaths. It is immediately relevant to monitor such simple instruments to assess humanity’s readiness to overcome the threat of climate change. The climate campaigner, Greta Thunberg, when asked by the US Congress on what to do on climate change, responded simply, “We want the science to be heard.” (*The Guardian*, October 2019).^20^ This tool is a simple instrument to monitor any groups’ readiness, including that of the US Congress, to tackle climate change.

In addition to illuminating such secular challenges, this work also speaks to practical steps when facing global threats. It demonstrates that digital technologies are effective even under severe and restrictive conditions of deployment. Leakages are inevitable, improvements are obvious - including to registration and monitoring - but digital cash transfer can reduce excess deaths worldwide as shown later. To my knowledge this has not been shown at such scale. Despite the digital divide between and within countries,^21^ the world has managed to deploy practical digital solutions and reap healthy digital dividends, i.e. lives saved. According to the WHO,^2,9^ excess deaths were 14.9 million and this work shows that the average treatment effect or semi-elasticity of the digital cash transfer is a statistically significant reduction of 1.53%.

To state the obvious, no countries acted on well written plans when the pandemic hit their borders. Equally, no ready-made formal economic theory is available to guide probability distributions of key unknowns in the relation I am investigating.

Therefore I offer this as an exploration rather than a definitive explanation. One thing is clear, not least to exclude reverse causality, all variables except digital cash transfer were measured before the pandemic (2014 – 2019). This then allows me to focus on the effect of digital cash transfer on excess deaths in 170 countries.

### Hypotheses and connections

To guide further analysis two hypotheses are tested. First, over other confounders such as the global north-south distinction, high trust in science is associated with fewer excess deaths. Moreover, digital cash transfers cause fewer excess deaths.

Two mechanisms connect digital cash transfer with risks of covid-19 deaths. First, through obviating the need for personal contact during civil-service-delivered support, digital payment reduces the required physical movement or mobility, thus reducing the risk of virus contractions. In contrast, non-digital payment during the delivery often involves personal contacts which increases the risks of virus contractions. Second, non-digital payment is also more susceptible to corruption which turns a meagre amount into a mere token, without any change in the risk of virus protection.

How could digital transfer/payment lead to fewer non-covid deaths, hence excess deaths? Take cancer or heart disease or any other non-communicable disease. The two connect indirectly through (a) hospital capacity and (b) forgone care. At times hospitals were overwhelmed with covid-19 patients during the pandemic. It follows that hospital capacity to treat non-covid patients (e.g. cancer patients) was severely reduced. Moreover, hospital doctors, nurses and other workers also contracted SARS-CoV2, pulling them away from the hospitals and clinics, which further diminished the capacity, this time across the board (serving covid and non-covid patients). Training specialist consultants, such as cancer surgeons or heart surgeons, to replenish this lost capacity would have taken longer than the three years of world emergency. All this contributed to excess deaths (covid-19 and non-covid-19). Incidentally, for some time in the future the capacity of health systems across the board will be lower than it would have been had the number of covid-19 patients been smaller. This calls for separate investigations.

Forgoing care is another link in how digital payment or its lack – which led to overwhelmed hospitals – can lead to excess deaths, especially among patients of non-communicable diseases. Seeing the overwhelmed hospitals and consuming the infodemic through social media (e.g. injecting disinfectant being an effective treatment of covid-19), these patients were understandably reluctant to go to hospitals for appointments.^22^ This effect was even greater among those with cancer and heart diseases because covid-19 death rates among patients with these pre-existing conditions increased considerably compared to those without them. Together, the supply side of hospital capacity and the demand side of patients’ reluctance combined to drive the increase in excess deaths once covid-19 threatened to overwhelm a country’s health system.

To preview the results, I found that digital cash transfers during the pandemic led to fewer excess deaths in 170 countries. Moreover, trust in science is found to be preventive. I also found that the global north reported lower excess deaths, no doubt partly due to the hoarding of vaccines early on in the pandemic which prompted the Director General of WHO to remind these countries about the global nature of the pandemic. Nowhere is safe unless everywhere is safe is the injunction that requires repeated reminders, reinforcing the enmeshed nature of our existence in the face of global challenges. A pandemic and climate change are among those challenges that can be better tackled using lessons offered here.

## MATERIALS AND METHODS

The outcome of excess death is entered in log form following the literature.^8,9,11^ The WHO furnished three sets of excess deaths figures: high, medium and low. It has assigned a few countries negative excess deaths.^2^ This means the log applied to these countries’ figures results in omitted outcomes. The are few negative excess death figures in the high set (4 out of 194 countries: China, New Zealand, Seychelles and Antigua) whereas for the other two sets there are up to 54 countries with negative excess deaths (more than one quarter of the world sample). Thus, the high set is chosen as the main model. Then a sensitivity analysis is conducted with the other two sets to test whether the omission of large number of countries makes a difference to the causal estimates obtained with the main or high set. Features of the WHO data will require care in use, ultimately requiring sensitivity analysis in any investigation. This is because most countries still do not have a system of civil registry and vital statistics that is working well in normal times, let alone in a pandemic.

Nevertheless, the release of these sets of excess deaths covering the world in such a timely manner is a milestone in the history of global public health. The earlier flu pandemic of 1918 received no concerted effort with countries hardest hit such as India obtaining death estimates much later and many countries were simply omitted. The WHO estimate therefore is invaluable information for its breadth, rigour, comparability and timeliness.

The causal variable, digital cash transfer, is drawn from the monitoring reports and database released by the World Bank.^5–7^ A country has digital cash transfer (versus non digital or no cash transfer) if it is listed as such in the database. A few words follow on the binary trust in science variable derived from the Wellcome Global Monitor 2018.^12^ The fact that trust in science is measured simply by asking people on a scale from one to four appears simple. But simplicity when effective is a virtue. The measure was fielded in 2018 and was not designed to be used in explaining covid-19 excess deaths. After all, none had seen the pandemic coming. Its effectiveness or external validity recommends itself compared to other options. Ji and colleagues proposed trust in government to explain covid-19 deaths and used a Bayesian model to derive the latent variable trust in government.^11^ In a sensitivity analysis I compare the two trust measures and their performance in explaining excess deaths. The simple construct of trust in science fits the data better. Any world survey should consider eliciting this information with the simple instrument.

The digital readiness/adoption index in 2014 was drawn from the database supplement of the World Development Report 2016 on *Digital Dividends*.^23^ The index summarises how ready the various sectors of the country (government, business, public) to adopt or create digital solutions in their operations. Higher index means more readiness to adopt digital solutions, and in this instance, it is likely to be correlated with the probability of adopting digital cash transfer during the pandemic. Other variables were drawn from the World Bank’s development indicators database including population size, percentage of older population (over 65 years), average income per person, percentage of mobile phone ownership and an indicator of high income economies.

### Endogenous treatment model

Log excess deaths is explained by digital cash transfer and trust in science across the world as follows:

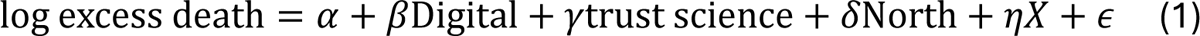

where all the three key exposures are binary variables: digital is whether the country used digital cash transfer,^5–7^ trust is whether the country has high (above median) trust in science^18,19^ and North (or high income countries according to the World Bank). The other confounding factors are included in the matrix *X*: average income per person, log population size, percentage of older population, percentage of mobile phone ownership whereas *X*_2_below includes digital readiness index in 2014 and indicator of global North.

But Digital is likely correlated with ε because of omitted variables i.e. there may be factors related to countries’ ability to deploy digital cash transfer which are also related to their performance in shaping public health outcomes. Thus Digital is endogenous. Two of the more recent Nobel prizewinners in economics suggest solutions, but these may not be applicable or credible in this instance. Randomised experiments for instance have been used in much recent research to estimate the causal effect of cash transfer; but randomising digital cash transfer during the pandemic, especially when vaccines were not yet developed, may be fraught with ethical challenges. It is no surprise that during the pandemic the AEA Registry received no randomised trials of digital cash transfer with excess deaths as the primary outcome (accessed 27 March 2024). Another solution to estimating the causal effect which has also been recognised with a Nobel prize is by resorting to natural experiments or instruments such as rainfall anomalies.^24^ In this instance there are hardly any credible natural experiments such as weather anomalies that can be used as instruments. The potentials or counterfactuals of not giving cash support during lockdown in the light of 7 million certified deaths due to covid-19 is bordering on the unconscionable. Therefore any modelling or analytic effort like this can only be seen as exploratory rather than definitive because such an answer when compared with a counterfactual as above is fraught and hardly credible. At least, it would be fraught with ethical challenges.

To overcome endogeneity and obtain the causal effect of digital cash transfer I therefore augment the main equation above with a second equation to explain adoption of digital cash transfer with a new variable from much earlier in time viz. digital readiness index in 2014. Digital cash transfer during the pandemic is explained by digital readiness index in 2014 as follows

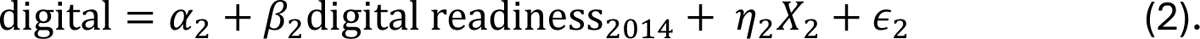

It is not assumed that digital readiness in 2014 is exogenous to the main/excess death equation – no assumption that ∈ ⊥ ∈_2_. Instead, because digital cash transfer is endogenous in the main equation, corr(∈, ∈_2_) ≠ 0. This is tested below. In the potential outcomes framework, once observed factors were considered, the difference between countries which adopted versus did not adopt is down to digital cash transfer alone.

## RESULTS

I draw a map of covid-19 excess deaths around the world, darkening the one-million country club which tallied excess deaths above the mark including India, Russia, Indonesia and the U.S.^8,9^

I then explore how much variations in log excess deaths relate to digital cash transfer and the two key factors of north-south divide and high-low trust in science by plotting their kernel densities in figure 2. The graphs strongly suggest that the log transform is warranted, though entailing omissions of four countries with negative excess deaths (China, Bhutan, Seychelles and Antigua). In defence of such omissions, analyses of alternative outcomes (high, medium and low sets) are done to check robustness below. Three features stand out in the data: unimodal, symmetric and almost smooth with default bandwidth.

**Figure 1.**
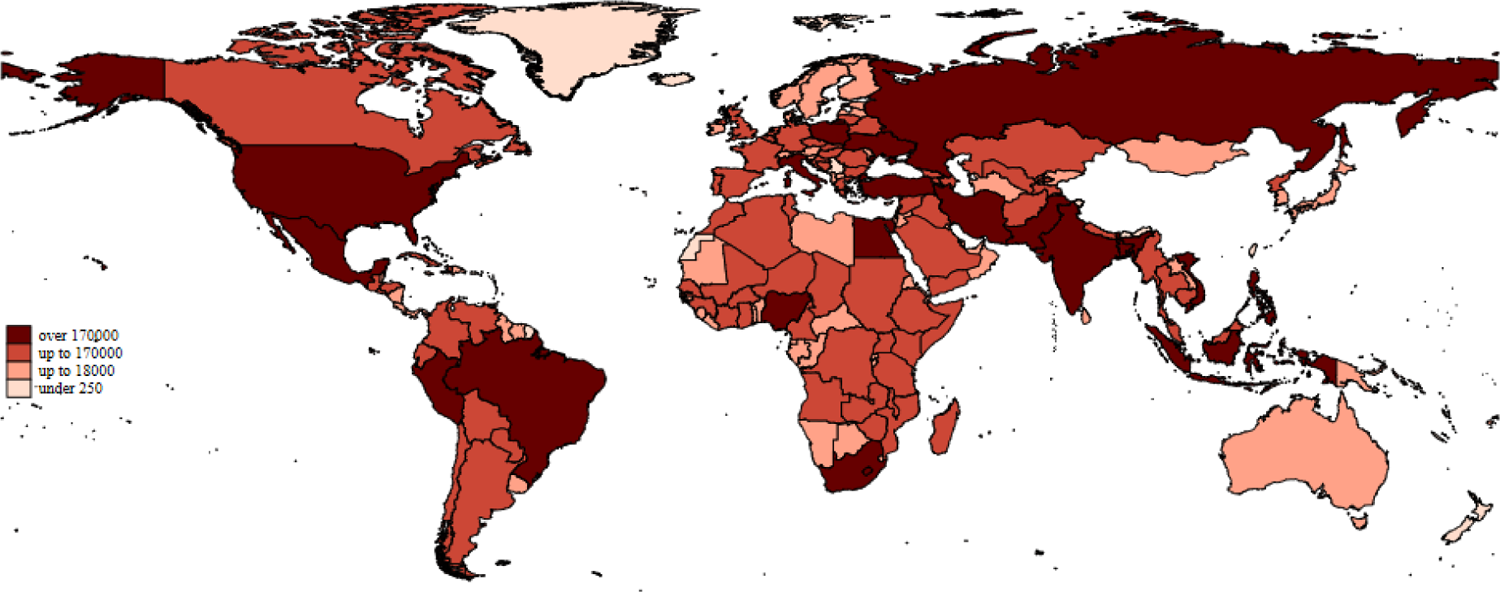
Map of covid-19 excess deaths around the world using WHO data, May 2023.

**Figure 2.**
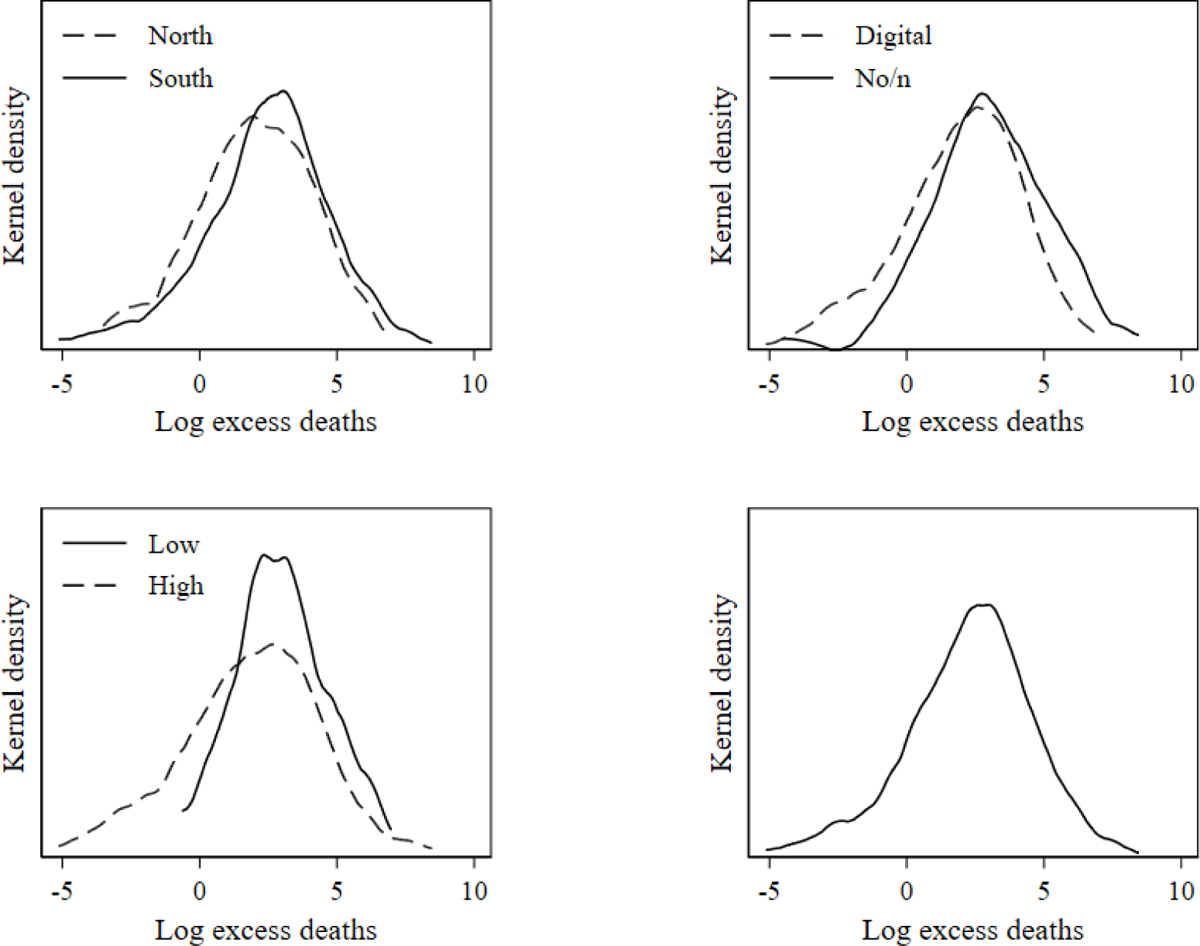
Kernel densities of log excess deaths by north – south (top-left), digital – non-digital cash transfer (top-right) and low – high trust in science (bottom-left).

The smoothed kernel densities in global north and global south (top-left pane) show that the density in the global south (solid line) is pushed right and up – there were more deaths in the global south and these were more concentrated and more precise in inference. Similarly, when comparing those of digital and non-digital cash transfer densities (top-right), that of non-digital cash transfer is pushed right and up – countries without digital cash transfer recorded more excess deaths. The same patterns are evident when low is compared to high (trust in science) with those countries with low trust in science recording more excess deaths. These are also more concentrated. Note that only one of these three variables is hypothesised as a cause viz. digital cash transfer (top right).

The analytic sample consists of 170 countries mostly from the south which also recorded higher numbers of excess deaths (log excess death on average 2.4 and standard deviation 2.4). Categorical variables are described in counts and proportions. We see in the continents of Asia, Africa (and Middle East) and South America there are more countries from the south whereas in rest of the world there are more countries from the north (Table 1). As recorded in the World Bank database not all countries in the north used digital cash transfer.^5–7^ Therefore a sensitivity analysis is called for, focusing only on countries in the south.

**Table 1.** Analytic observations for explaining log excess deaths from covid-19 around the world.

|  | South<br>(N=119) | North<br>(N=55) | Total<br>(N=170) |
| --- | --- | --- | --- |
| Log excess death/1000 | 2.4 (2.4) | 2.0 (2.2) | 2.3 (2.3) |
| Digital cash transfer |  |  |  |
| No or non-digital | 72 (60.5%) | 50 (90.9%) | 122 (70.1%) |
| Digital payment | 47 (39.5%) | 5 (9.1%) | 52 (29.9%) |
| Trust in science 2018 |  |  |  |
| Low | 63 (52.9%) | 5 (9.1%) | 68 (39.1%) |
| High | 56 (47.1%) | 50 (90.9%) | 106 (60.9%) |
| Continent |  |  |  |
| Others | 26 (21.8%) | 39 (70.9%) | 65 (37.4%) |
| Asia | 27 (22.7%) | 6 (10.9%) | 33 (19.0%) |
| Africa, Middle East | 49 (41.2%) | 7 (12.7%) | 56 (32.2%) |
| South America | 17 (14.3%) | 3 (5.5%) | 20 (11.5%) |
| Log population 2019 | 16.0 (2.1) | 15.5 (1.9) | 15.8 (2.1) |
| Percent older population 2019 | 6.2 (4.1) | 15.1 (6.4) | 9.0 (6.5) |
| Mobile phone per 100 people 2019 | 98.4 (34.7) | 129.1 (22.4) | 108.1 (34.4) |
| log average income 2019 | 8.8 (0.9) | 10.7 (0.4) | 9.4 (1.2) |
| Digital readiness 2014 | 0.4 (0.1) | 0.7 (0.1) | 0.5 (0.2) |

The first table of estimates (table 2) shows three sets of associations with log excess deaths (high, medium and low sets). The three sets differ in the number of observations reflecting the different number of countries with negative excess deaths recorded in the WHO database. The high set has the largest sample.

**Table 2.** Log excess deaths associations around the world using three sets of WHO estimates: high (170 countries), medium (165) and low(122).

|  | Log excess<br>death,<br>High (N=170) | Log excess<br>death,<br>Medium (165) | Log excess<br>death,<br>Low (122) |
| --- | --- | --- | --- |
| Digital transfer | -0.283**<br>(-2.22) | -0.397**<br>(-2.03) | -0.630**<br>(-2.40) |
| Global north | -0.922***<br>(-4.96) | -1.153***<br>(-4.07) | -1.558***<br>(-4.48) |
| High trust in science | -0.244**<br>(-2.05) | -0.350*<br>(-1.94) | -0.388<br>(-1.62) |
| Log population | 1.082***<br>(36.05) | 1.132***<br>(24.26) | 1.090***<br>(16.85) |
| Pensionage2019 | 0.00434<br>(0.30) | -0.0213<br>(-0.96) | 0.0423<br>(1.41) |
| Mobile phone2019 | 0.00340<br>(1.65) | 0.00601*<br>(1.93) | 0.00485<br>(1.16) |
| Log income2019 | 0.00968<br>(0.11) | 0.265**<br>(2.00) | 0.540***<br>(2.69) |
| Region (ref: rest of world) |  |  |  |
| Asia | -1.243***<br>(-6.51) | -1.663***<br>(-5.60) | -0.765*<br>(-1.72) |
| Africa, Middle East | -0.768***<br>(-4.08) | -1.263***<br>(-4.44) | -0.638<br>(-1.52) |
| South America | -0.259<br>(-1.30) | -0.276<br>(-0.90) | 0.443<br>(1.09) |

|  | adj. R2 | 0.91 | 0.83 | 0.78 |
| --- | --- | --- | --- | --- |
| *t* statistics in parentheses |  |  |  |  |
| * $p < 0.1$, ** $p < 0.05$, *** $p < 0.01$ | | | | |

The three columns of estimates show two important patterns. First, irrespective of the number of omitted countries (due to negative excess deaths), digital cash transfer is associated with fewer excess deaths – and it is always statistically significant (first row). The results therefore are not sensitive to the omissions. Second, the other two key factors, whose smoothed densities are shown above, show similarly significant associations, with one exception. Note that all these associations are not causal, because digital cash transfer may be endogenous which will be addressed later. Trust in science is also associated with fewer excess deaths and significantly so except for the low set (right column). This non-significance is the result of the small number of observations (one quarter less than the high set). The last in the three key factors viz. north-south also shows an expected association where the north has significantly fewer excess deaths. There are variations across regions of the world that are often significant (Asia reported fewer than Europe or North America). Population size, not proportion of older people, is the more significant demographic factor. This is not surprising, because the outcome is not only covid-19 deaths (which are prevalent in the older population) but also indirectly related deaths (general population).

The next estimates of causal effect on log excess deaths (main equation, Table 3) are obtained using an endogenous treatment model, where digital cash transfer is in turn explained by the digital readiness index in 2014 (the second equation). Moreover, a correlation (testing for endogeneity) between the error terms is also estimated.

**Table 3.** Effect of digital cash transfer on log excess deaths using three sets of WHO estimates: high (170 countries), medium (165) and low (122).

|  | Log excess death, High (N=170) | Log excess death, Medium (165) | Log excess death, Low (122) |
| --- | --- | --- | --- |
| Main |  |  |  |
| Digital transfer | -1.533***<br>(-8.43) | -2.175***<br>(-6.87) | -2.152***<br>(-5.06) |
| North | -1.319***<br>(-6.02) | -1.726***<br>(-5.34) | -1.914***<br>(-5.39) |
| Trust science | -0.246***<br>(-2.72) | -0.287**<br>(-2.18) | -0.409**<br>(-2.30) |
| Log population | 1.093***<br>(40.88) | 1.143***<br>(27.18) | 1.115***<br>(19.80) |
| Pension age 2019 | 0.0160<br>(1.10) | 0.00350<br>(0.10) | 0.0492**<br>(2.11) |
| Mobile phone 2019 | 0.00327**<br>(2.17) | 0.00628**<br>(2.57) | 0.00531<br>(1.35) |
| Log income 2019 | 0.00112<br>(0.01) | 0.218<br>(1.57) | 0.447*<br>(1.90) |
| Region (ref: rest of world) |  |  |  |
| Asia | -1.164***<br>(-6.06) | -1.275***<br>(-3.42) | -0.708*<br>(-1.84) |
| Africa, Middle East | -0.640***<br>(-3.55) | -1.034***<br>(-3.09) | -0.644*<br>(-1.94) |
| South America | -0.178<br>(-1.11) | -0.0280<br>(-0.11) | 0.495*<br>(1.65) |
| Second (digital transfer) |  |  |  |
| Digital readiness 2014 | 1.913**<br>(2.00) | 2.103<br>(1.62) | 3.892**<br>(2.56) |
| Log income 2019 | -0.261<br>(-1.47) | -0.316<br>(-1.41) | -0.766***<br>(-2.74) |
| North | -1.249***<br>(-4.15) | -1.138***<br>(-3.89) | -0.943***<br>(-2.59) |
| var(logexcess) | 0.680***<br>(6.98) | 1.471***<br>(3.72) | 1.436***<br>(4.54) |
| corr(transfer,logexcess) | 0.927***<br>(26.60) | 0.915***<br>(18.61) | 0.788***<br>(8.22) |
| adj. R2 | 0.78 | 0.76 | 0.76 |

*t* statistics in parentheses
\* $p < 0.1$ , \*\* $p < 0.05$ , \*\*\* $p < 0.01$

The focus is on the first column which has the largest number of countries (170), though the results are robust to omissions of countries with negative excess deaths (columns 2 and 3). Digital cash transfer reduces excess deaths during the pandemic and this effect is highly significant (*p*<0.001). The semi-elasticity or average treatment effect of digital cash transfer is 1.53%. The smaller samples give larger average treatment effects with the same significance, therefore the model in focus i.e. the high set (left column) gives an average treatment effect that is most conservative. Continuing with the other two key factors (north-south and trust in science), these are also highly significant and remain negatively associated with excess deaths. The magnitudes, however, are telling. Since all are binary variables, it is possible to gain some sense of comparison (a more definitive comparison will be plotted later). The effect of digital cash transfer is largest, even compared to the association with north-south divide (−1.533 versus −1.319). Trust in science has a smaller coefficient though it is also significant.

The correlations between the errors from the main and digital cash transfer equations range from 0.788 to 0.927 (highly significant), suggesting that digital cash transfer is endogenous, which required care in estimation such as applied here. And briefly, the other covariates are similar to the model earlier, which did not account for endogeneity. Again, this is consistent across the three analytic samples which vary in size, suggesting robust results. As expected, digital readiness in 2014 correlates with digital cash transfer six years later. The average potential outcomes for the two counterfactuals are shown in table 4 which gives the average treatment effect of −1.533.

**Table 4.** Potential outcomes in two counterfactuals.

|  | Margin | Std. Err. | z | P | Conf. | Interval |
| --- | --- | --- | --- | --- | --- | --- |
| No or non-digital | 3.123 | .081 | 38.36 | <0.001 | 2.964 | 3.283 |
| Digital | 1.591 | .135 | 11.75 | <0.001 | 1.326 | 1.857 |

To focus on comparing the key risk factors (trust in science and north-south divide) and the cause (digital cash transfer), I plot their marginal effects/associations below (Figure 3; no confidence intervals are shown in order to obtain clear plots – the significance in all marginal plots is referenced to table 3 above). The plots show that trust in science is associated with fewer excess deaths but the magnitude of the association is small compared to the causal effect of digital cash transfer. However, the north-south divide association is nearly as large as the causal effect of the digital cash transfer. This is consistent with the early actions of the global north to retain as much vaccine as possible for their populations, delaying distributions to the global south.

**Figure 3.**
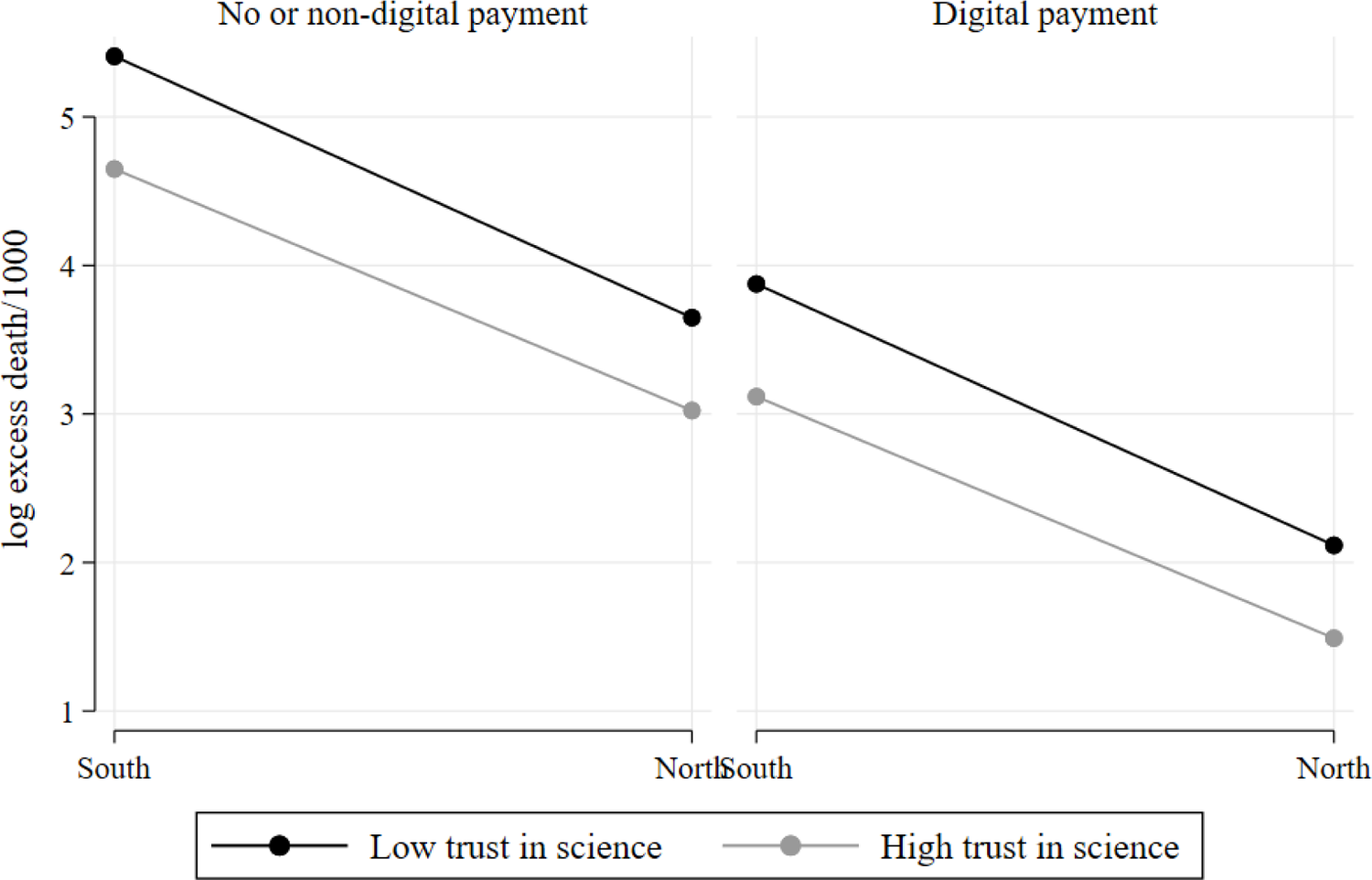
Plots of effects of digital cash transfer compared with those of trust in science and of South-North divide; in each plot differences across (e.g. South and North) are not causal, differences down are causal.

The digital cash transfer makes a considerable difference to any combination of the key associations. The plot shows that the worst combination is in the South with low trust in science and no digital cash transfer, while the best is in the North with high trust in science and digital cash transfer.

### Heterogeneity

As mentioned above there maybe heterogeneity between the North and South so the endogenous treatment model is re-estimated separately on the sample with the most countries (table 5). The heterogeneity is only hinted at because focusing on the global south reduces the sample size drastically, widening confidence intervals. The endogenous treatment model gives comparable size −0.240 though this is no longer significant. Estimating two equations simultaneously on a much reduced sample may give imprecise estimates. More importantly, the residual correlation between the two (equations’) error terms is no longer significant (last row, standard error 0.927) suggesting that the adoption of digital cash transfer can be treated as exogenous. If this is followed up, we get the results in the second column. Doing so or when the focus is on association, digital cash transfer is significantly related to fewer excess deaths and significant (coefficient −0.291). Together these estimates support the idea that the main results are robust.

**Table 5.** Heterogeneity: in the south.

|  | Log excess<br>death,<br>High (N=170) | Log excess<br>death,<br>High (N=170) |
| --- | --- | --- |
| Main |  |  |
| Log population | 1.074***<br>(0.000) | 1.075***<br>(0.000) |
| Pensionage2019 | 0.047*<br>(0.020) | 0.048*<br>(0.019) |
| Mobile phone2019 | 0.000<br>(0.890) | 0.000<br>(0.868) |
| Log income2019 | 0.084<br>(0.349) | 0.081<br>(0.359) |
| Region (ref: rest of world) |  |  |
| Asia | -0.735***<br>(0.000) | -0.738***<br>(0.000) |
| Africa, Middle East | -0.447*<br>(0.011) | -0.447*<br>(0.016) |
| South America | -0.127<br>(0.488) | -0.125<br>(0.512) |
| Trust science | -0.166<br>(0.101) | -0.166<br>(0.119) |
| Digital transfer | -0.240<br>(0.668) | -0.291*<br>(0.013) |
| Constant | -14.751***<br>(0.000) | -14.728***<br>(0.000) |
| Digital transfer |  |  |
| Digital readiness | 3.521*<br>(0.018) |  |
| Log income2019 | -0.405<br>(0.074) |  |
| Constant | 1.929<br>(0.220) |  |
| var(logexcess) | 0.256*** |  |
|  | (0.000) |  |
| corr(transfer,logexcess) | -0.063 |  |
|  | (0.927) |  |
| N | 118 | 118 |

*p*-values in parentheses
\* $p < 0.05$ , \*\* $p < 0.01$ , \*\*\* $p < 0.001$

### Two mechanisms: contingent social contract and weakening informality

Eventually, widespread vaccine coverage effectively controlled the pandemic, but initially there was no certainty that efficacious vaccines would be developed in time. Until then, governments imposed social restrictions or non-pharmaceutical interventions of various kinds and strengths. Based on what pandemic history, especially the flu pandemic of 1918, teaches that social restrictions were associated with fewer deaths, it is through these restrictions that the digital cash transfer realises its effect in lowering excess deaths. Evidence in support of these mechanisms is shown in table 6, demonstrating that digital cash transfer is strongly and significantly associated with more stringent restrictions. This illustrates how governments and societies are involved in social contract bargaining, where in return for societies acceding to government imposed restrictions, society members receive government cash transfer by digital means. The two key means to eventually defeat covid-19 and return societies to normal activities, namely vaccines and restrictions are effective in combination, the latter helped by digital cash transfer.

**Table 6.** Stringency index during the pandemic (Source: Oxford data) and digital cash transfer.

|  | Stringency,<br>N=159 |
| --- | --- |
| Log population | 1.302***<br>(2.64) |
| Pensionage2019 | -0.486**<br>(-2.14) |
| Mobile phone2019 | 0.00535<br>(0.16) |
| Log income2019 | 7.206***<br>(5.18) |
| Region (ref: rest of world) |  |
| Asia | 1.694<br>(0.58) |
| Africa, Middle East | -1.261<br>(-0.41) |
| South America | 1.033<br>(0.32) |
| North | -6.729**<br>(-2.28) |
| Trust science | 2.262<br>(1.21) |
| Digital transfer | 4.315**<br>(2.22) |
| adj. R2 | 0.280 |

One group of these restrictions is readily available to any government, unlike the other one of vaccine development. For most countries in the world rapid vaccine innovation to tackle rapidly mutating viruses is beyond their capacity. Erecting restrictions, on the other hand, lies firmly within government purview. With digital cash transfer in place governments can credibly tighten restrictions as part of the government-society social contract. Given digital cash transfer, citizens find it easier to comply with more stringent restrictions, hence these become more effective. This contract between credible government and compliant citizens translates to the magnitude and statistical significance of the digital cash transfer variable.

While the above mechanism applies around the world, the second mechanism operates in the south viz. through weakening the correlation between informality and excess deaths. For this exercise I constructed a factor of informality rates from the ILO and the World Bank,^25^ retaining the first factor scores. This is because informality rates are likely measured inconsistently and intermittently across countries in the global south, giving an erroneous variable in any one source. When there are multiple sources of measurements, factor analysis is typically used to obtain a better measure of informality.^26,27^

The results in table 7 are telling. The first column shows that informality is associated with more excess deaths (coefficient 0.067), though not statistically significant – perhaps due to the very small sample size, less than half of the main table. When digital cash transfer is included, two things happened. Consistent with the main results (table 3), digital cash transfer is associated with fewer excess deaths and significantly so, plus the coefficient of informality is almost halved (0.038). Together these associations illustrate the mechanism by which the digital cash transfer weakens the link between high rates of informality and high rates of deaths from covid-19.

**Table 7.**
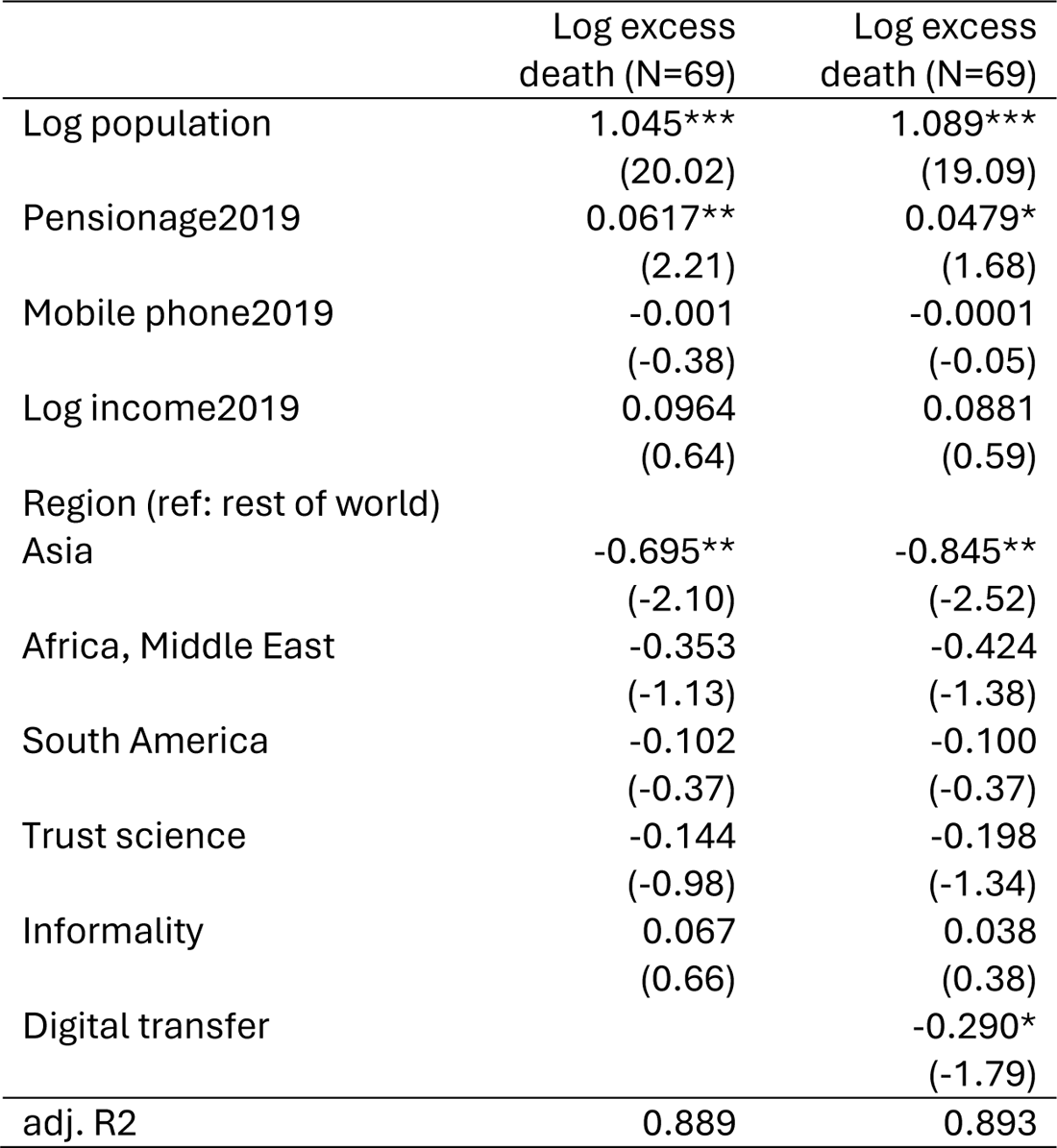
Stringency index during the pandemic (Source: Oxford data) and digital cash transfer.

|  | Log excess<br>death (N=69) | Log excess<br>death (N=69) |
| --- | --- | --- |
| Log population | 1.045***<br>(20.02) | 1.089***<br>(19.09) |
| Pensionage2019 | 0.0617**<br>(2.21) | 0.0479*<br>(1.68) |
| Mobile phone2019 | -0.001<br>(-0.38) | -0.0001<br>(-0.05) |
| Log income2019 | 0.0964<br>(0.64) | 0.0881<br>(0.59) |
| Region (ref: rest of world) |  |  |
| Asia | -0.695**<br>(-2.10) | -0.845**<br>(-2.52) |
| Africa, Middle East | -0.353<br>(-1.13) | -0.424<br>(-1.38) |
| South America | -0.102<br>(-0.37) | -0.100<br>(-0.37) |
| Trust science | -0.144<br>(-0.98) | -0.198<br>(-1.34) |
| Informality | 0.067<br>(0.66) | 0.038<br>(0.38) |
| Digital transfer |  | -0.290*<br>(-1.79) |
| adj. R2 | 0.889 | 0.893 |

### Sensitivity analyses

One set of sensitivity analyses was conducted throughout i.e. using the other two sets of excess deaths released by the WHO (altogether working with the high, medium and low sets of figures). The main tables above (2 and 3) show that the omission of some countries does not materially affect the causal effect of digital cash transfer. In fact, the high set gives the most conservative causal effect. Neither did the omission affect the associations with north-south divide and trust in science.

In this way, this work fits in the recent literature which has explored various forms of trust and their associations with deaths from covid-19. Trust in government has been found to associate with fewer deaths due to covid-19.^11^ The authors have kindly made their data available for examination. It is likely that such trust in government is also associated with the outcome of *excess* deaths, for the same direct and indirect connections laid out above. One set of sensitivity analysis is to compare trust in government and trust in science – side by side. See table 8 where standardised coefficients are shown (note varying number of observations).

**Table 8.**
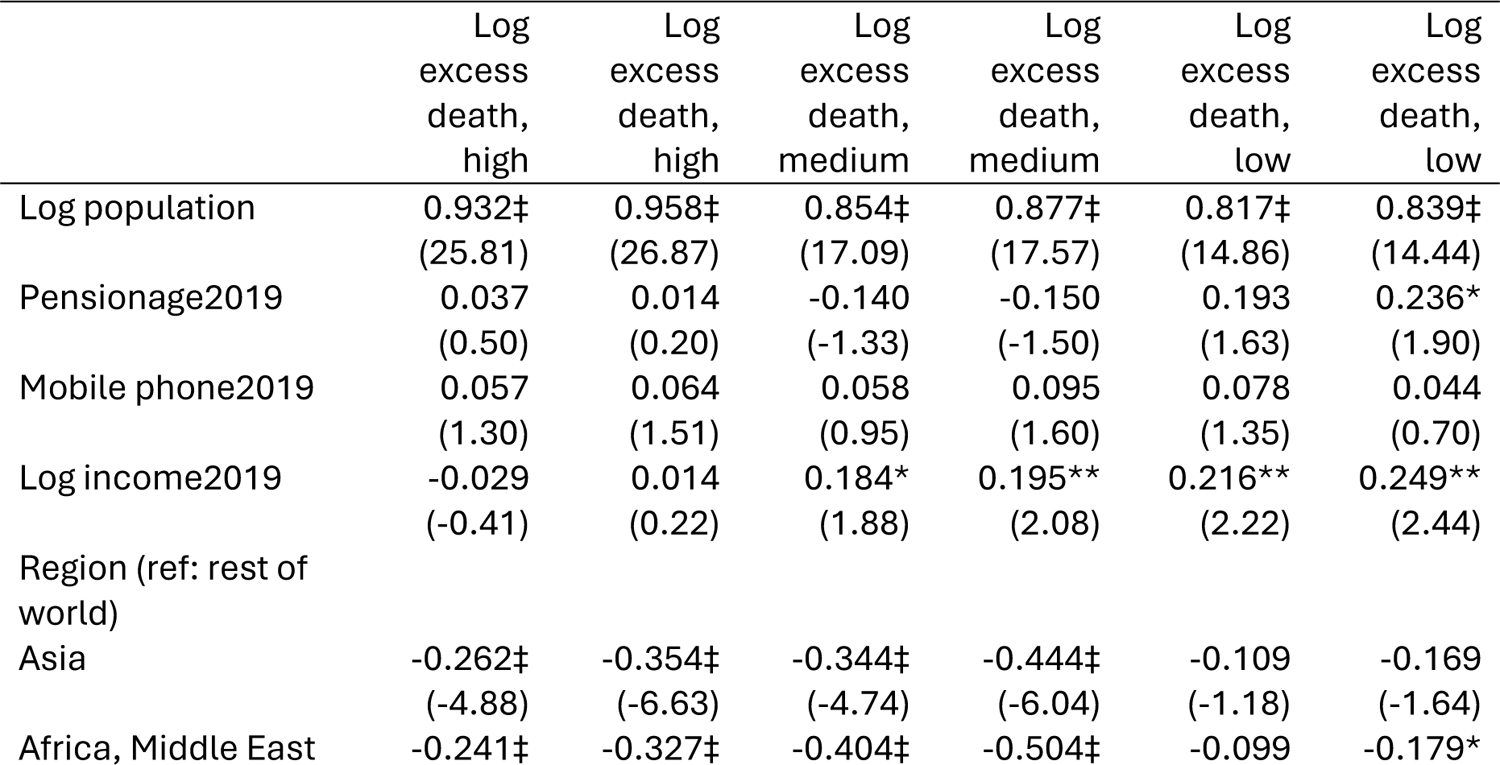
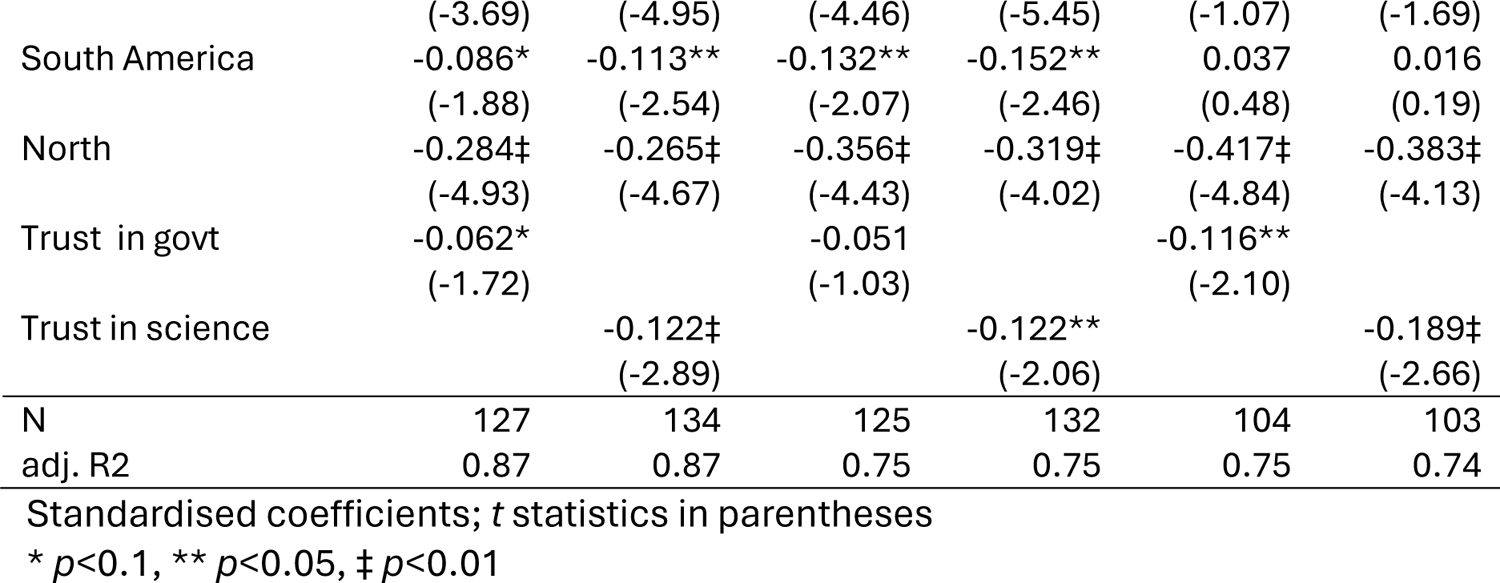
Comparing trust in government and trust in science in associations with log excess deaths, showing standardised coefficients for comparison.

Both kinds of trust are statistically significant and negatively associated with excess deaths, except once (middle column) where trust in government is not statistically significant. Both trusts are raw scores to enable standardised coefficients to be compared. At the foot of the table are fit statistics which suggest that both trusts fit the data equally well. But there are differences. The standardised coefficients of trust in science are larger in magnitude - for each standard deviation change in trust in science the absolute magnitude is considerable. Now, the same standardised coefficient of trust in government also associates with excess deaths, but to a lesser extent. This sensitivity analysis suggests that excess deaths is better explained by trust in science than trust in government. Moreover, the stronger association with trust in science also suggest a leverage for intervention.

These results can be further examined, specifically by including both types of trust to gain further appreciation of which one explains better the world’s experience throughout the pandemic. The results are collected in table 9 which shows that trust in science remains significant and negative while trust in government is no longer significant. Together, both tables suggest that trust in science on its own better explains excess deaths from covid-19, supporting the main model specification which omits trust in government.

**Table 9.** Comparing trust in government (Ji et al 2024) and trust in science, together, in associations with log excess deaths, showing standardised coefficients for comparison.

|  | Log excess death, high | Log excess death, medium | Log excess death, low |
| --- | --- | --- | --- |
| Log population | 0.950***<br>(24.50) | 0.871***<br>(15.82) | 0.833***<br>(13.83) |
| Pension age 2019 | 0.014<br>(0.17) | -0.185<br>(-1.60) | 0.206<br>(1.54) |
| Mobile phone 2019 | 0.043<br>(0.90) | 0.057<br>(0.84) | 0.069<br>(1.08) |
| Log income 2019 | -0.012<br>(-0.16) | 0.198*<br>(1.85) | 0.221**<br>(2.13) |
| Region (ref: rest of world) |  |  |  |
| Asia | -0.326***<br>(-5.56) | -0.424***<br>(-5.24) | -0.148<br>(-1.40) |
| Africa, Middle East | -0.310***<br>(-4.41) | -0.497***<br>(-4.99) | -0.149<br>(-1.43) |
| South America | -0.122**<br>(-2.49) | -0.178**<br>(-2.55) | 0.013<br>(0.15) |
| North | -0.243***<br>(-3.84) | -0.304***<br>(-3.41) | -0.364***<br>(-3.69) |
| Trust in govt | -0.043<br>(-1.10) | -0.019<br>(-0.33) | -0.071<br>(-1.09) |
| Trust in science | -0.120**<br>(-2.52) | -0.134*<br>(-1.93) | -0.145*<br>(-1.79) |
| N | 118 | 116 | 97 |
| adj. R2 | 0.87 | 0.74 | 0.74 |

## DISCUSSION: SERENDIPITY, CREATIVITY AND INEQUITY

This work set out to estimate – during the devastating covid-19 pandemic – how much difference digital cash transfer makes while exploring the role of trust in science.

Serendipity and creativity combined to make a huge difference. Trust in science, which the world serendipitously reported a few years before the pandemic, is associated with fewer excess deaths some years later. While acknowledging serendipity, creativity in deploying support through digital cash is even more important. Digital cash transfer reduces excess deaths considerably. This work demonstrates that digital cash transfer has reduced excess deaths, thereby impacting the expansion of life, a fundamental idea of development as an expansion of freedoms.^28,29^

This study is offered not as a definitive impact evaluation but an attempt or a base study of digital cash transfer effect. No doubt more research will examine the question of linking various forms of public support and deaths during the pandemic.

Given the sudden shock of the pandemic a binary variable of digital transfer constructed in the recent literature may be deemed inadequate.^5–7^ In their defence two things can be said. First, no randomised trial was registered during the pandemic with a precisely defined digital cash transfer binary variable with excess deaths as the primary outcome. It would be very hard to agree on the amount of transfer for nearly 200 countries around the world. Thus to avoid the precision paralysis which would have resulted had we insisted on a randomised binary treatment, one must proceed pragmatically. In doing so I focused on compiling variables available prior to the pandemic to avoid reverse causality.

What does this all mean in lives saved? Trust in science may have inoculated society against deaths during the pandemic. More importantly, by comparing the two coefficients of trust in science and digital cash transfers, it is found that the transfers have saved more lives. While these numbers are large and important their significance goes beyond this history of the global devastation brought by the novel virus SARS-CoV2. Two are particularly salient: climate change and digital dividend.

To return to Oreskes’ quote, we mistrust science at our peril as a human race living under a changing climate.^16^ The evidence here suggests that trust in science may have serendipitously inoculated the world against a shock of novel virulent pathogens. It would thus be unwise to distrust science when the world is facing the threat of climate change. The final clause of Oreskes above is apt. When the US Congress asked the climate change campaigner Greta Thunberg, she exhorted the American representatives to trust science.^20^ This is not a glib saying. On this side of the Atlantic a senior member of the government is famous for his riposte that the country had too many experts, implying their science should no longer be trusted.^17^ If one accepts the importance of trust in science (distinct from science itself) then this calls for efforts to monitor such trust globally in addition to efforts to support the sciences of these challenges e.g. global health or climate change. In practice, the simple instrument used here has proven its worth and is fit for the task of monitoring.

The value of this lesson on monitoring trust in science when facing global challenges such as climate change is made clearer by the similarity and difference between the pandemic and climate change. Both challenges have a quality of practical externality – none is safe until everyone is safe or all earth inhabitants live under a warming planet. The difference is during the pandemic, a country’s air space can be closed to prevent infection spreading (witness New Zealand). But no comparable tool – no air space to confine carbon dioxide to – is available to prevent climate change. This adds a critical need to trust in science to respond to climate change. Now, if indeed trust in science has a role to play in climate change response, I would hypothesise for future research that a country’s willingness to support restitution for ‘loss and damage’ due to climate change will be higher when its trust in science is also high.

### Digital cash transfer, healthy digital dividends and AI revolution

If trust in science traces a secular and long-term change, digital cash transfer was immediate and responsive. And it bore fruits of healthy digital dividends. The healthy digital dividend revealed here - healthy in the sense of life (not death) and in the sense of ample - is only one kind of digital dividend. Another kind that is widely discussed at the moment arises from the application of artificial intelligence and machine learning in medicine.^30^ The intensive work continues on applying these technologies to challenges posed by cancer, dementia and other devastating diseases. These can reduce “health inequity … if managed carefully” (Bill Gates, philanthropist and founder of Microsoft).^31^

Digital cash transfer technology and artificial intelligence are mutually symbiotic. Digital cash transfer laid out the data infrastructure which is the sine-qua-non for many applications of artificial intelligence and machine learning. In the artificial intelligence literature this is known as the ‘unreasonable effectiveness of data’ theorem.^32^ The healthy digital dividend revealed here helps pave the way for making the future AI revolution possible through the construction of its data infrastructure. In fact, we might have been there already as a suggestive piece of evidence has been put forward recently. By using advanced estimator in 146 countries, it shows that digital cash transfer can bring those outside the formal financial system into the fold. The digital cash transfer has broadened financial inclusion by 2% to 8%.^33^

Meanwhile, the literature on healthy digital dividends has been growing irrespective of the pandemic. The healthy digital dividend on the global scale reported here is not the first. In normal times digital technologies have been deployed and are saving lives. Ample literature has detailed how digital technologies have delivered development and expanded freedoms to be and do things we value.^23^ Most of these demonstrations arise in context that are general or prevalent such as in fishing communities and in rural areas facing threats from communicable diseases such as heart disease and diabetes.^34,35^ Some works in rural Indonesia before and through the pandemic have demonstrated this digital dividend in preventing deaths from heart disease throughout the pandemic.^35–37^ Though the pandemic is unique, it still offers general lessons on healthy digital dividend.

### Global health inequity

While serendipity and creativity are full of promise, a stark global health inequity is uncovered in this work. The global north recorded fewer excess deaths, even considering its disadvantageous population structure facing the virus SARS-CoV2. The north-south coefficient is significantly negative and sizeable. The unequal distribution of vaccines along the north-south divide prompted the head of the WHO to repeatedly remind the world that no one is safe as long as there are large reservoirs of covid-19 cases anywhere in the world, breeding randomly mutated variants of interest. No doubt more will be unearthed by scholarship in this area of covid-19 vaccines, their efficacies and their inequitable distribution.^38^

### Prior literature

Given the recency of the pandemic the literature is still growing, particularly on estimating the magnitude of excess deaths and on estimating risk factors associated with excess death variations across countries. The Covid-19 Excess Mortality Collaboration emphasises that it is important to assemble excess death figures for all countries despite two-thirds of countries around the world lacking death registration system.^8^ This is because excess mortality is fundamental to making decisions in public health (:1513). Thus the collaboration proceeded on the basis of reports from 74 countries to estimate and furnish excess death figures for 191 countries. Motivated by the same reason, the WHO proceeded to furnish updated sets of excess death figures for 200 countries. The Collaboration estimated 18.2 million whereas the WHO estimated 14.9 million excess deaths. This discrepancy can arise from the sheer shock to all health systems around the world (including in rich countries like the US which is in the million country club), thus strengthening the importance of explaining the causes behind the excess deaths.

Explaining this mortality statistic, particularly its variation across countries and its associated risk factors which are potentially modifiable, is the aim of the related literature which is also growing. Sun and colleagues studied factors associated with excess deaths in 80 countries and found six important variables.^10^ They distinguished explanatory variables in general into causes and associated factors, and concentrated on the latter. This is a useful distinction, especially since there is some overlap with the factors used here. Nevertheless there is a novel risk factor here i.e. trust in science which is sourced from the same survey they used, namely the Wellcome Global Monitor survey 2018.^12^ Notably it has been used before in an exploration of wealth inequality’s association with trust in science.^19^ Its construction is the same as in subsequent literature on trust and covid-19.^18^ Together these show that trust in science has some external validity. This work extends the literature not only in geography to 170 countries but also substantively as it analyses both cause and risk factors. Importantly, this work uses those factors available before the pandemic to avoid reverse causality.

### It is as well to note some limitations

Although this work deals with the endogeneity of digital cash transfer, it does not deal with the endogeneity of trust - the other key variable. This however is similar to other works.^11^ Even so it depends on assuming and testing for the residual correlation, not randomised trial. A rejoinder would be: it is hardly ethical to run a randomised trial when life and death are the stakes and when no one knew effective vaccines would come along. The causal effect obtained here, a step up from associations, is among the best estimates under the pandemic circumstance. Another limitation is possible measurement error especially on the outcome. Last, little doubt with time more data will become available especially those that are comparable across 200 or so countries, e.g. on vaccine hesitancy.^22,39^ It is likely that with such data at hand, they can enrich and not overturn the importance of both digital cash transfer and trust in science.

This work has three key strengths. It is the first to study excess deaths 170 countries, an important statistic for global health and health systems strengthening. Next, it does so with a parsimonious model. The parsimony arises partly from the fact that these variables are available, not because they are the most accurate. But this is common. Johns Hopkins and Oxford gave the same caveat viz. collecting from what were available from government sources with varying quality and quantity. Nevertheless the WHO and the World Bank are as good as any for sources on covid-19 and cash transfers. Last, the trust variable has previously shown some external validity.^2,5–7,9^

In conclusion, the investigation of excess deaths from covid-19 offers positive and negative lessons, ones that cannot be understood by studying only the period since China reported to the WHO cases of novel respiratory infections in Wuhan. Only by taking an extended view would the world be better prepared for global challenges. What happened years before the pandemic had material bearing on how many survived through it. First, trust in science is a precious global commodity irrespective of immediate threats to the world. Monitoring it at regular intervals seems wise. Second, even in a pandemic the world has been able to harness digital technologies to deliver healthy digital dividends. Third, a negative lesson must be heeded: health inequity along the north-south divide remains entrenched. But the technology and knowledge are there to make progress towards delivering health equity for all everywhere.

## Data Availability

All data produced in the present work are contained in the manuscript.

https://www.who.int/data/sets/global-excess-deaths-associated-with-covid-19-modelled-estimates

